# Evaluating ten commercially-available SARS-CoV-2 rapid serological tests using the STARD (Standards for Reporting of Diagnostic Accuracy Studies) method

**DOI:** 10.1101/2020.09.10.20192260

**Authors:** Laurent Dortet, Jean-Baptiste Ronat, Christelle Vauloup-Fellous, Céline Langendorf, David-Alexis Mendels, Cécile Emeraud, Saoussen Oueslati, Delphine Girlich, Anthony Chauvin, Ali Afdjei, Sandrine Bernabeu, Samuel le Pape, Rim Kallala, Alice Rochard, Celine Verstuyft, Nicolas Fortineau, Anne-Marie Roque-Afonso, Thierry Naas

## Abstract

Numerous SARS-CoV-2 rapid serological tests have been developed, but their accuracy has usually been assessed using very few samples, and rigorous comparisons between these tests are scarce. In this study, we evaluated and compared 10 commercially-available SARS-CoV2 rapid serological tests using the STARD methodology (Standards for Reporting of Diagnostic Accuracy Studies). 250 sera from 159 PCR-confirmed SARS-CoV-2 patients (collected from 0 to 32 days after onset of symptoms) were tested with rapid serological tests. Control sera (N=254) were retrieved from pre-COVID periods from patients with other coronavirus infections (N=11), positive rheumatoid factors (N=3), IgG/IgM hyperglobulinemia (N=9), malaria (n=5), or no documented viral infection (N=226). All samples were tested using rapid lateral flow immunoassays (LFIA) from ten manufacturers. Only four tests achieved ≥98% specificity, with other tests ranging from 75.7%-99.2%. Sensitivities varied by the day of sample collection, from 31.7%-55.4% (Days 0-9), 65.9%92.9% (Days 10-14), and 81.0%-95.2% (>14 days) after the onset of symptoms, respectively. Only three tests evaluated met French Health Authorities’ thresholds for SARS-CoV-2 serological tests (≥90% sensitivity + ≥98% specificity). Overall, the performances between tests varied greatly, with only a third meeting acceptable specificity and sensitivity thresholds. Knowing the analytical performance of these tests will allow clinicians to use them with more confidence, could help determine the general population’s immunological status, and may diagnose some patients with false-negative RT-PCR results.

## INTRODUCTION

Asymptomatic carriage of SARS-CoV-2 has been estimated in some studies to be as high as 86 (1). Others posit that it may be responsible for up to two-thirds of viral propagation (1-4). As the world increasingly acknowledges the challenges this poses to disease containment, reliable testing has become central to monitoring the COVID-19 pandemic, informing health policy, rapidly responding to events as they evolve, and mitigating disease transmission (5, 6).

Yet, RT-PCR tests (Real-time reverse transcription-polymerase chain reaction), the gold standard for SARS-CoV-2 detection, have substantial limitations. PCR requires specialized, expensive laboratory equipment, is often only located in laboratories with biosafety level ≥2, and may be affected by sample transport delays of 2-3 days, in which time COVID-19 suspects may further expose other patients and health workers (7-9). For SARS-CoV-2, RTPCR testing also uses naso-pharyngeal swab samples that can be complex to obtain, pose considerable risk to health care workers with insufficient personal protective equipment (PPE), and produce false-negative results in up to 30 of confirmed COVID-19 patients (10-12). Chest radiography (CXR) and computed tomography (CT) scans are currently used to overcome PCR tests’ lack of sensitivity but also require expensive equipment (11, 13). These challenges limit current molecular and imaging approaches’ ability to be scaled up in epidemic settings where rapid, reliable, and easy population screening is needed.

Thus, serological confirmation of COVID-19 antibodies could provide an important complementary tool to PCR testing by identifying previously exposed individuals (8, 12). SARS-CoV-2 seroconversion occurs 7-14 days after the onset of symptoms (8, 14-16). A few classic ELISA tests (enzyme-linked immunosorbent assays) are currently available, but considerable effort has been made by manufacturers to offer even earlier rapid diagnostic tests (RDTs) (17). According to the Foundation for Innovative New Diagnostics (FIND), 177 SARS-CoV-2 antibody RDTs were commercially-available on June 15^th^, 2020 (18). Most informations were directly submitted by test suppliers or obtained from publicly available sources and were not independently verified. Neither their analytical performance nor their usefulness in a clinical setting has yet been rigorously evaluated with a sufficient panel of samples (19, 20). In addition, validation criteria seem to be different from one country to another (21-23).

We carried out a retrospective clinical evaluation of ten commercially available RDTs, comparing their performance, according to the delay between the onset of symptoms and sampling, severity of the disease and usability of the tests. Our study was designed using the 2015 Standards for Reporting of Diagnostic Accuracy Studies (STARD) (24). We aim to provide accurate clinical performance data to assess the RDTs’ utility and their ability to be integrated into adapted diagnostic algorithms across health systems and epidemiological contexts, especially in areas with limited resources (24).

## METHODS

### Study design

We conducted a retrospective study on 250 serum samples collected between March 11 till April 3^rd^ from 159 patients, 256 sera with documented RT-PCR positive results for SARSCoV-2 using nasopharyngeal swabs (eSwabs™-Virocult, Copan, Italy). Real-time RT-PCR targeting RNA-dependent RNA polymerase and E genes were used to detect the presence of SARS-CoV-2 as described by Corman *et al*. (7).. All patients were from 2 University hospitals located in the south of Paris (Bicêtre and Paul Brousse Hospitals) and provided between one and four serum samples. Sera from COVID-19 patients were randomly selected and grouped according to the time between onset of symptoms and patient’s blood sampling (0-9 days, 10-14 days, and > 14 days) (Fig. 1A).

**Figure 1.**
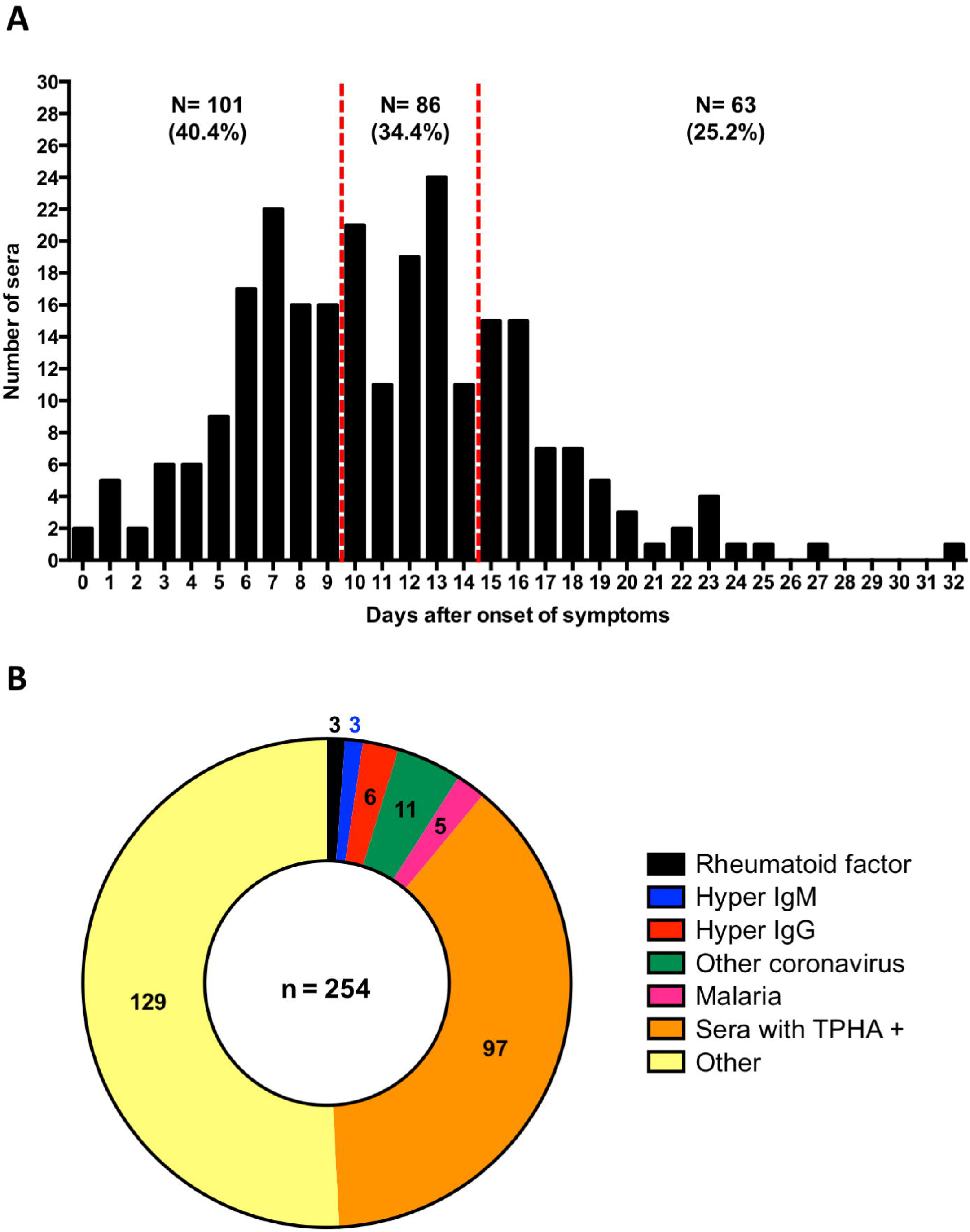
Sera collection used for the evaluation. A. Distribution of 250 sera from COVID positive patients according to the number of days after onset of symptoms. B. Distribution of the 254 control sera.

To assess specificity, an additional 254 sera collected prior to December 2019 were selected, and which had previously been tested positive for a separate agent or pathology that could potentially interfere with SARS-CoV-2 testing results, either other coronavirus (n=11), other viral infections (n= 129), a rheumatoid factor (n=3), hyperglobulinemia IgG (n=6) and IgM (n=3), malaria (n=5), or a *Treponema pallidum* hemagglutination assay (TPHA) (n=97) (Fig. 1B).

Each RDT was evaluated on the same collection of sera. The minimum sample size was calculated assuming an expected sensitivity of 90 (with 5 accuracy) and a specificity of 98 (with 2 accuracy), amounting to 250 true positive samples and 254 true negative samples (power 0.80, alpha 0.05).

### Sample preparation

Selected sera were randomly placed in working boxes so as not to bias technicians’ interpretation of results. Two sets of these boxes were prepared and stored at 4°C prior to being used.

### Selected Tests

Diagnostic tests were selected based on supply, expected performance (based on published literature), and on commercial brochures. Ten RDTs that could detect either all antibodies or specifically identified IgG or IgM (in blood, serum, or plasma) were evaluated: (RDT 1) NG-Test IgG-IgM COVID-19 (NG-Biotech, Guipry, France), (RDT 2) Anti SARS-CoV-2 rapid test (Autobio Diagnostic CO, Zhengzhou, China), (RDT 3) Novel Coronavirus -2019-nCOV-Antibody IgG/IgM (Avioq Bio-tech CO, Yantai, China), (RDT 4) NADAL^®^ COVID-19 IgG/IgM Test (Nal Von Minden GmBH, Regensburg, Germany), (RDT 5) Biosynex^®^COVID-19 BSS (Biosynex, Illkirch-Graffenstaden, France), (RDT 6) 2019-nCoV Ab Test (Innovita Biological Technology CO, Qian’an, China), (RDT 7) 2019-nCoV IgG/IgM (Biolidics, Mapex, Singapore), (RDT 8) COVID-19-CHECK-1 (Veda.Lab, Alençon, France), (RDT 9) Finecare SARS-CoV2 Antibody test (Guangzhou Wondfo biotech, Guangzhou, China) and (RDT 10) Wondfo SARS-CoV2 Antibody test (Guangzhou Wondfo biotech, Guangzhou, China). Characteristics of these RDTs are summarized in Table S1. Tests were performed at room temperature by trained laboratory technicians. All tests followed the manufacturers’ instructions, strict biosecurity measures, and good microbiological practices and procedures [8].

The intensity of the reaction line was recorded in 3 gradations: No signal (0), very weak but definitively positive (1), and medium to high intensity (2). Values were not recorded when a control line did not appear, when tests would subsequently be repeated (Fig. S1A and B).

Visual test interpretation was conducted independently by two separate readers and recorded on data collection sheets. Readings were determined based on two of three readers’ interpretations. In cases where all three interpretations were different; results were registered as unknown.

### Data analysis

Each RDT’s sensitivity and specificity was calculated with its respective confidence interval 95 (CI95) using VassarStats (http://http://vassarstats.net/).

The cumulative positivity at different points of illness (from symptom appearance until day 31 post-appearance) was determined as follows (i) a positive result on Day N was followed by subsequent positive results on Days N+1, N+2, N-n, etc and (ii) a negative result on Day N was preceded by negative results on Days N-1, N-2, N-n, etc. Details of the calculation are presented in Figure S2.

Cumulative curves were fitted to an asymmetrical (five-parameter) logistic equation using Graph Prism v6 (25). For comparative purposes, the point at which 50 cumulative positivity was reached was calculated for all RDTs and expressed as the number of days post-symptom onset (Fig. S3, Table 1).

**Table 1.** Median times for SARS-CoV-2 seroconversion using 10 commercially available RDTs, Paris, France, June 2020.

| RDT | Median time to seroconversion |  |
| --- | --- | --- |
|  | Days after symptom onset | CI95 |
| 1 | 8.3 | (8.2 - 8.4) |
| 2 | 7.4 | (7.3 - 7.6) |
| 3 | 7.0 | (6.8 - 7.1) |
| 4 | 7.2 | (7.0 - 7.3) |
| 5 | 7.8 | (7.6 – 7.9) |
| 6 | 9.6 | (9.5 - 9.7) |
| 7 | 8.2 | (8.1 – 8.4) |
| 8 | 7.5 | (7.4 - 7.7) |
| 9 | 7.0 | (6.8 - 7.1) |
| 10 | 7.0 | (6.8 - 7.1) |

CI95, 95 confidence interval

The positive predictive value (PPV) and negative predictive value (NPV) were calculated as follows: PPV = (sensitivity x prevalence) / [(sensitivity x prevalence) + ((1 – specificity) x (1 – prevalence))], and NPV = (specificity x (1 – prevalence)) / [(specificity x (1 – prevalence)) + ((1 – sensitivity) x prevalence)].

### Usability Evaluation

A self-administered user experience questionnaire using the Osgood scale was used for all tests and focused on the clarity of the instructions for the test user, the test’s technical complexity, the ease of test result interpretation, and access to legal information (26).

### Ethics

All samples were from a Bio-bank (BIOCOVID-19) after having received ethical clearance from the Patient Protection Committee (PPC) of the Ile-de-France VII (No. 2009-965). Blood samples from patients infected with the SARS-CoV-2 virus, who were subjected to routine testing as part of clinical management but whose serum samples had not been entirely used for clinical purposes, were approved for use in this study. The biobank is stored in CRB Paris South (BRIF: BB-0033-00089). The planning, conduct, and reporting of studies was in line with the Declaration of Helsinki.

## RESULTS

### Clinical characteristics of COVID-19 patients

Overall, 250 sera collected from 159 COVID-19 patients were selected from the BIOCOVID19 Bio-bank. The median age was 62.9 years (range 12.8 - 97.6) and the male/female ratio was 1.69 (100/59). Among these individuals, 4.4 (7/159) were discharged after their initial visit to the emergency room (ER) and 95.6 (152/159) were hospitalized. Over the study period, 44.1 (67/152) of patients required ICU care while hospitalized. The overall death rate among hospitalized patients was 19.1 (29/152); 10.5 (9/85) among non-ICU patients and 29.9 (20/67) among ICU patients. Most sera were sampled on Days 0-15 (85.5, 219/256) after symptoms appeared, though sera from later dates (up to Day 31) were also available (Fig. 1A).

### Test Performance

Cumulative positivity rate rose with time, reaching 100 at 20-days post-symptom onset for all RDTs (Fig. 2). More than 50 of SARS-CoV-2 infected patients had detectable antibodies 7 to 10 days after symptoms appeared (Fig. 2). The time needed to reach >95 sensitivity varied between 14 days (for half of the RDTs tested) and 18 days (for RDT 6) (Fig. 2). Asymmetrical (five-parameter) logistic analysis demonstrated that 50 cumulative positivity (or the median time for seroconversion) varied from 7.0 to 9.6 days (Table 1).

**Figure 2.**
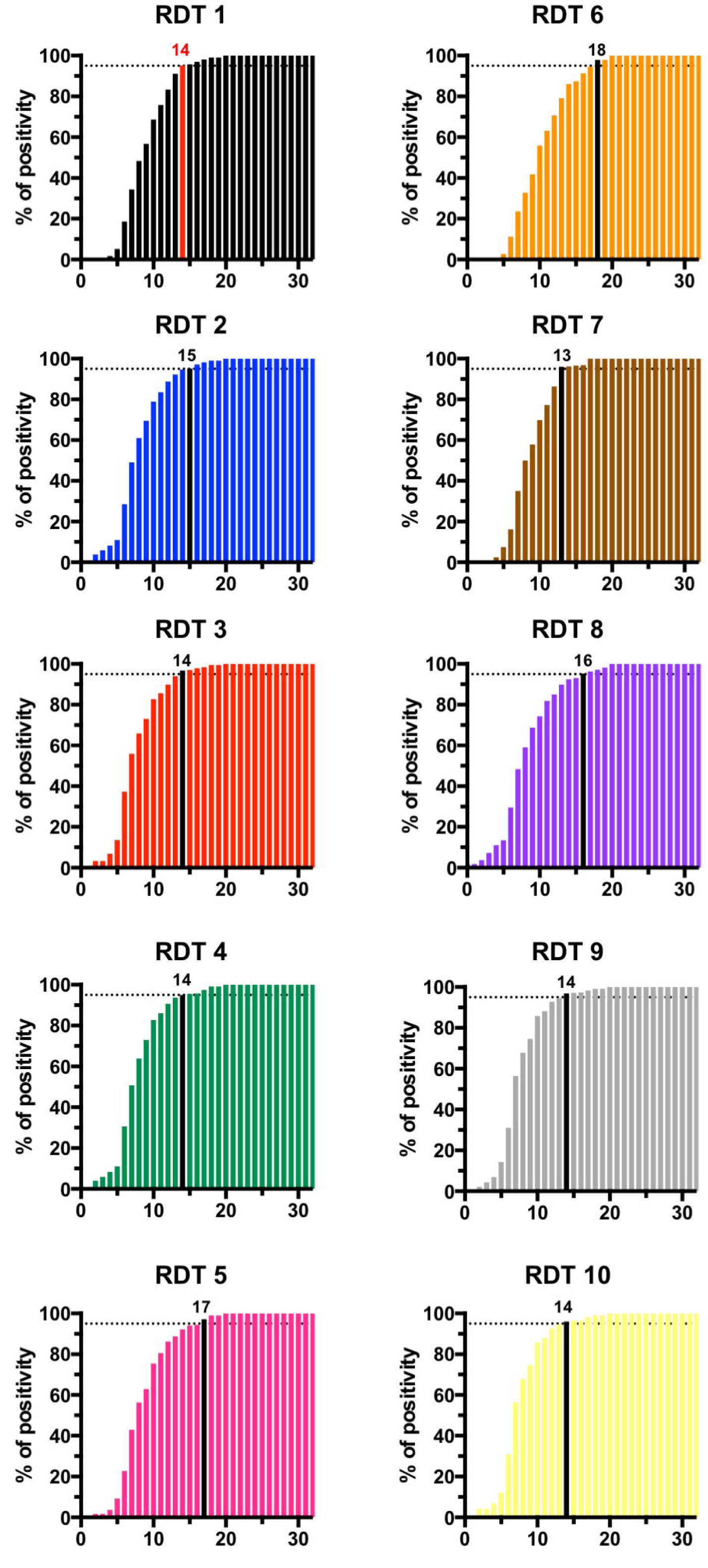
Cumulative positivity rate obtained with 10 RDTs in sera from COVID-19 patients stratified by the number of days after appearance of symptoms. The day after symptom appearance with >95 positivity is indicated by a coloured bar (red for RDT 1, black for the other tests)

As expected, overall test sensitivity was highest 15 days after the appearance of symptoms (Table 2), with all RDTs reaching >90 sensitivity at that point, except for RDT 6 and RDT 8 (81.0 and 88.5, respectively). For the 8 RDTs able to differentiate between IgM and IgG, combined detection significantly increased overall test sensitivity with the exception of RDT 1, RDT 4 and RDT 5 (for which IgM detection seemed to be nearly as sensitive as IgM + IgG detection) (Table 2).

**Table 2.**
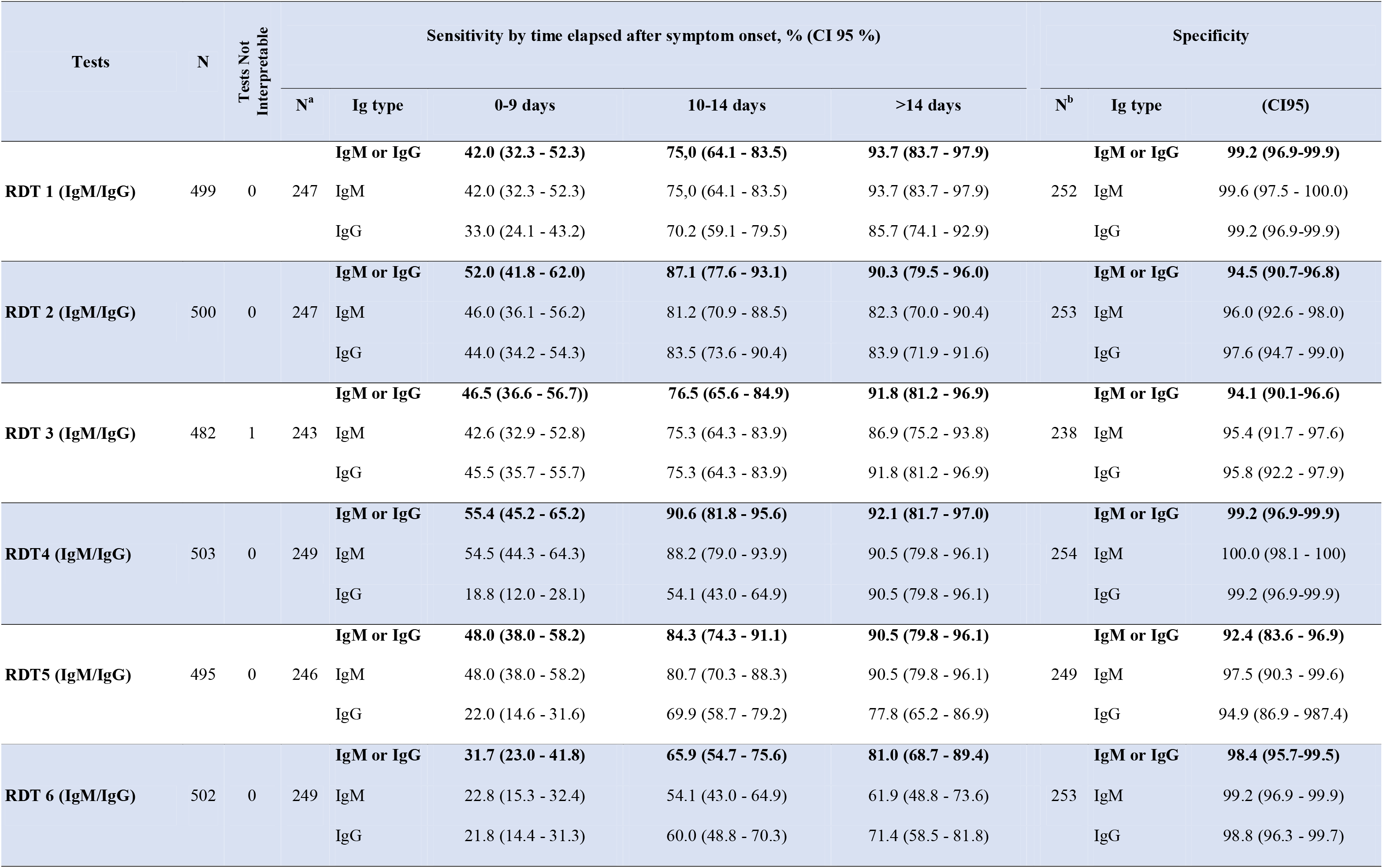
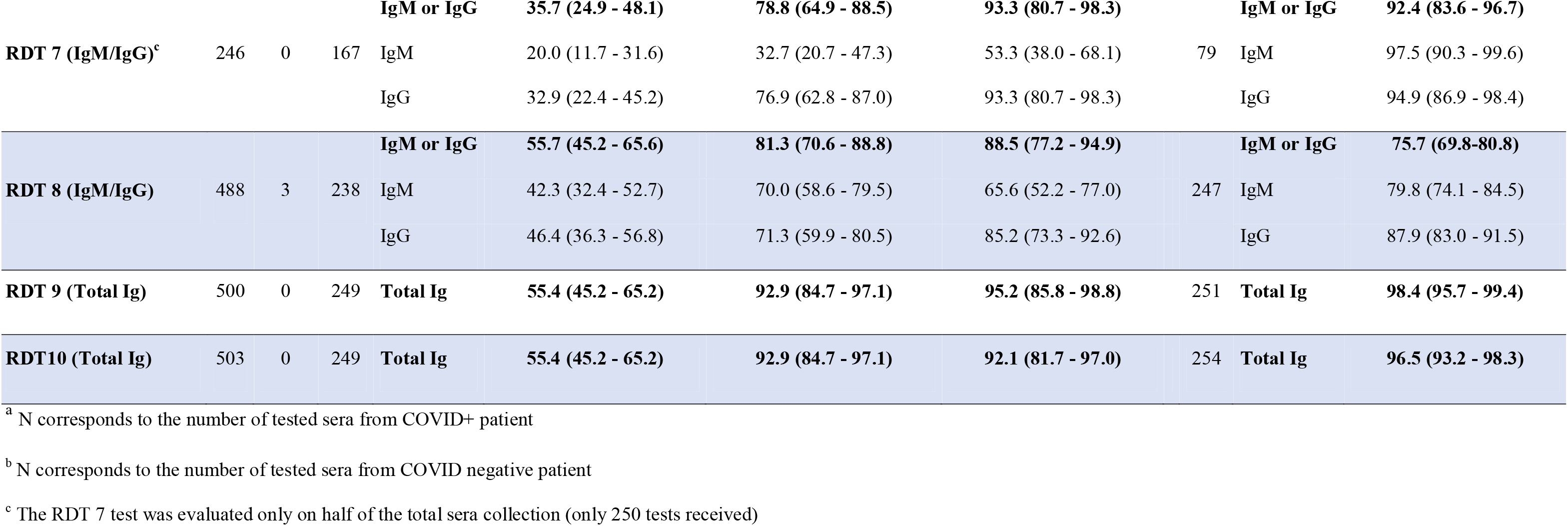
Performance of 10 rapid serological tests for SARS-CoV-2 antibodies, Paris, France, June 2020.

Specificities, calculated with sera recovered from patients between 2017 and early 2019, ranged from 75.7 to 99.2. Only four tests (RDT 1, RDT 4, RDT 5 and RDT 9), reached the >98 threshold recommended by the French Health Authorities for serological diagnostic tests (Table 2) (23). The presence of a rheumatoid factor did not induce false positive results except in the case of the RDT 3, which systematically gave a positive IgM (3/3) and/or IgG (1/3) signal. Among the 11 sera with a non-SARS-CoV-2 agent (other coronaviruses) four tests produced one false positive result and one test produced two false positives. Notably, the false positives occurring in non-SARS-CoV-2 agent samples corresponded to one serum recovered from the same patient. No other patterns were detected for other false positive results (Table S2). The concordance between all tests varied from 77.0 to 96.4 except in the case of the RDT 8 test that had a lower concordance with other RDTs (<80). Other RDTs gave concordant results (usually ~90 to 95) (Fig. 3).

**Figure 3.**
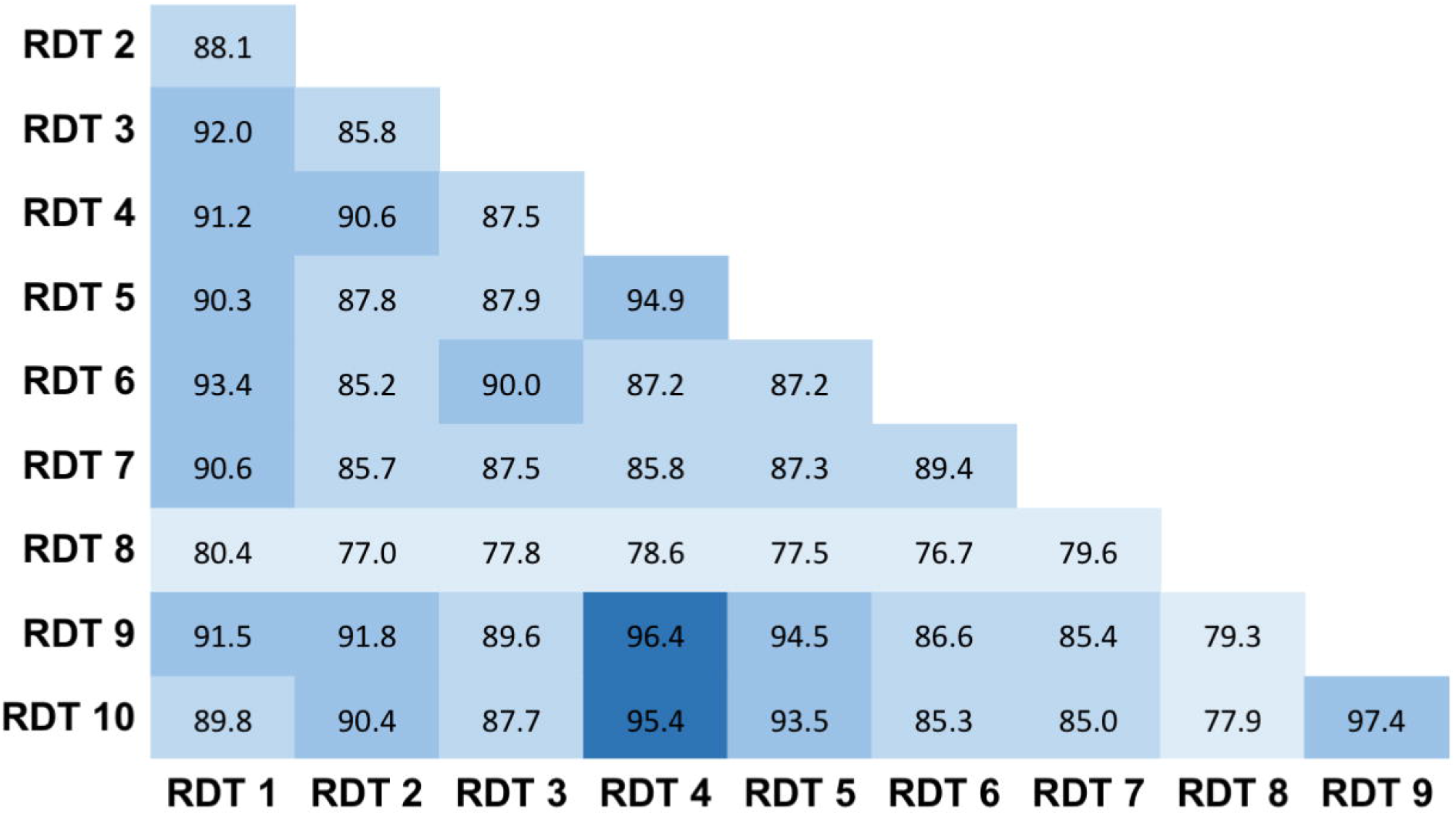
Results agreement between RDTs. Percent agreement is indicated across all RDT combinations. RDTs were considered positive if any of IgG and/or IgM was detected.

The positive and negative predictive values (PPV and NPV respectively) describe the performance of a diagnostic test. A high result can be interpreted as indicating the accuracy of such a test. The PPV and NPV are not intrinsic to the test (as true positive rate and true negative rate are) but they depend also on the prevalence. As the prevalence increases, the PPV also increases but the NPV decreases. Similarly, as the prevalence decreases the PPV decreases while the NPV increases. As a consequence, having both VPN and PPV above a certain value can be quite challenging. Among the 10 RDTs evaluated only three presented PPV and NPVs above 95 over a large window of population prevalence (RDT1, RDT 4, and RDT9) (Fig. S4).

### Band intensity

To compare the ease of reading the RDTs’ banded results, the intensity of the reaction line was recorded according to 3 gradations: No signal (0), very weak but definitively positive (1), and medium to high intensity (2). As shown in Figure 4, the overall ease of reading was highest for sera recovered >14-days after the appearance of symptoms. Band intensity was most prominent in tests with combined antibody detection (i.e. both IgM and IgG detection; RDT 9 and RDT 10 tests) (Fig. 4A). Among the eight RDTs that differentiated between antibody types, IgM band intensity was most pronounced with RDT 1 test (Fig. 4B), with RDT 3, RDT 4 and RDT 5 tests closely following. Conversely, IgM bands obtained with the RDT 6, RDT 7, RDT 8 and, to a lesser extent, the RDT 2 tests were significantly less pronounced (Fig.e 4B). For IgG tests, the bands produced by the RDT 1, RDT 2, RDT 3, RDT 7 and RDT 8 tests were more prominent than the RDT 4, RDT 5 and RDT 6 tests (Fig. 4C).

**Figure 4.**
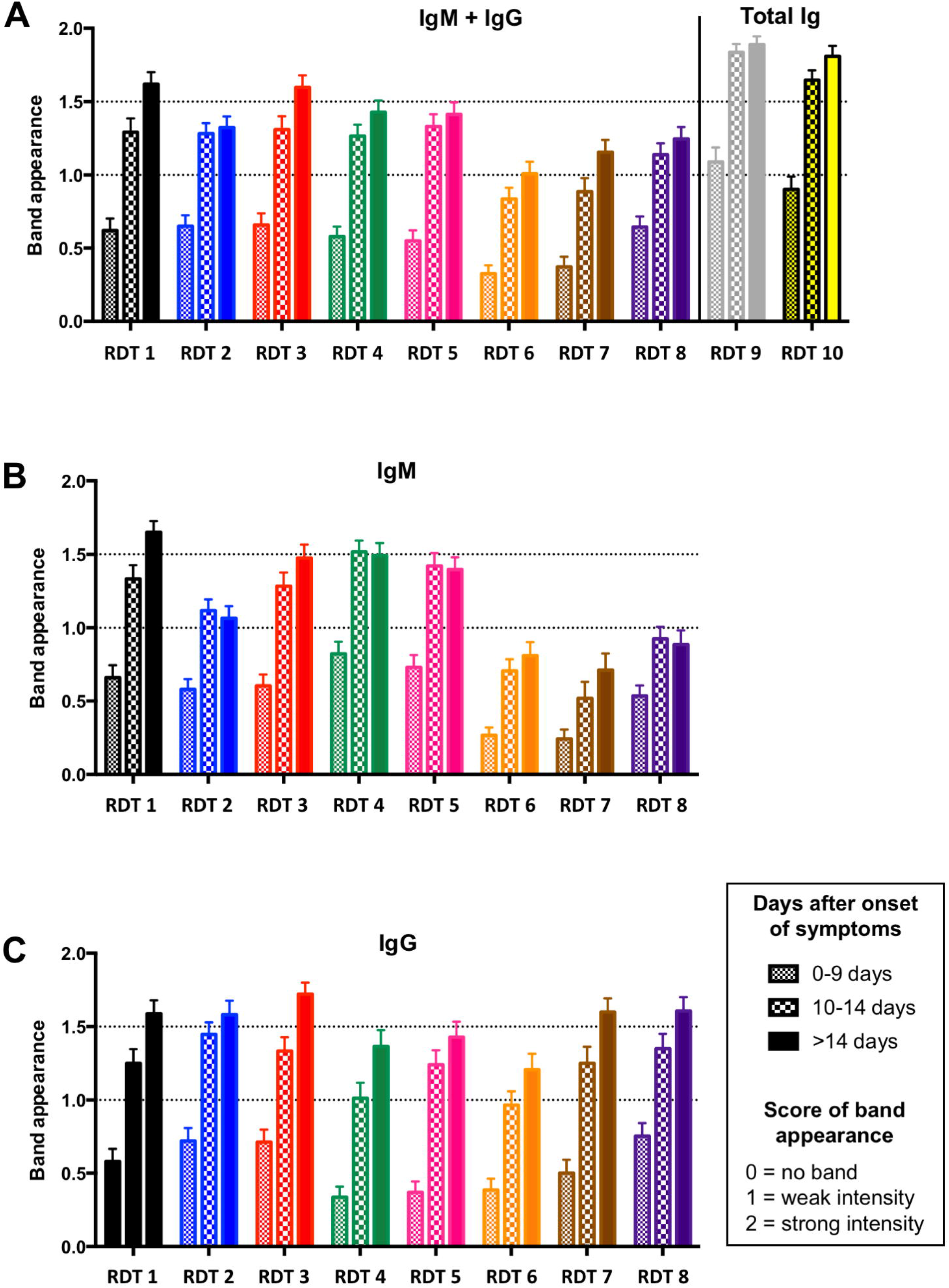
Results (visible band) intensity for IgM + IgG (panel A), IgM only (panel B), and IgG only (panel C) tests.

### Ease-of-Use

All the tests were in cassette form and nearly all devices used standard colloidal gold antigen conjugated particles (Table S1). One test (RDT 9) used fluorescent antigen conjugated particles for visualisation using a specific reader. Ease-of-use could vary from one test to another, and all contained ‘Instructions For User’ (IFU) manuals that were in all cases considered easy to understand (Table S3). Only RDT 9’s IFU did not provide figures explaining their methods or results interpretation. Most (6/10) IFUs contained figures explaining their methods and results interpretation, and 3/10 IFUs contained figures explaining results interpretation (Table S3). No users reported difficulty using the RDTs, though the RDT 2 test provided a dropper with no clear instruction as to how many drops should be used. Buffer for RDT 9 was included with every test tube. Less than half of the RDTs (RDT 1, RDT 3, RDT 4, and RDT 5) included single-use plastic pipettes or similar devices for transferring samples into the test wells. No users reported difficulties identifying sample and buffer wells. All tests’ results interpretation, with the exception of RDT 6, were considered easy. The recommended time-to-read results ranged from 10-20 minutes (Table S1). From a packaging and legal point of view, all manufacturers except RDT 6 respected the CE-IVD regulation to describe needed storage conditions in the IFU, on test packaging, and in product references. RDT 6’s reference test was not found on the box nor within the IFU. All tests were in a single, sealed package and included a desiccant pouch.

## DISCUSSION

With no curative medications currently available for COVID-19 and vaccines in early stages of development, social distancing and widespread testing have become the primary tools available to control an unprecedented global health crisis. Serological assays and RDTs are being increasingly used across the world to address other tests’ limitations, but most commercially available RDTs have had their accuracy verified on only a small number of sera without including negative samples to evaluate cross-reactivity. Moreover, their usefulness for patient management in active hospital settings and among the general public has almost never been rigorously evaluated (27, 28). By demonstrating the feasibility and accuracy of rapid serological immunoassays with a substantially more robust sample size than has previously been described, we add depth to the evolving conversation surrounding SARSCoV-2 testing strategies. We hope that knowing the analytical performance of nearly a dozen commercially available tests, and by providing comparative detail, we will allow clinicians to select and use these tests with more confidence and certainty.

This study is, to our knowledge, the first to compare diagnostic performance and time-toseropositivity in nearly a dozen SAR-CoV-2 RDTs using a large sample size (250 selected samples each for specificity and sensitivity, more than double other peer-reviewed, published RDT evaluations). Other studies evaluating antibody tests have also not included samples from patients with non-SARS-CoV-2 infections to evaluate specificity.

Overall, after the appearance of symptoms, seroconversion occurred on Days 7-9 for 50 of COVID positive patients (Table 1), with >95 seroconverting after 14 days using RDT 1, RDT3, RDT 4, RDT 9 and RDT 10) and 18 days for RDT 6) (Fig. 2). The specificities ranged from 94.5-99.2, except for RDT 8 test (75.7). Notably, the RDT 3 test produced systematic false positive results with sera of patients who had a high level of rhumatoid factor (Table S2).

Thresholds for sensitivity and specificity for RDTs have been set by many National Health Authorities (21-23). For diagnosis in symptomatic patients, high sensitivity is required (generally ≥90), while specificity is less critical as some false-positives may be tolerated as other potential diagnoses are considered in parallel (RT-PCR and/or CT scans). However, if LFIAs were deployed as an individual-level approach to inform release from quarantine or immune-protection, then high specificity (>98) is essential, as false-positive results return non-immune individuals to risk of exposure (23). Using the French health authority [21] acceptable limits for SARS-CoV-2 serological tests (≥90 sensitivity; ≥98 specificity) our evaluation validated only three RDTs for clinical use, namely NG-Test IgG-IgM COVID-19 (RDT 1, NG-Biotech), NADAL^®^ COVID-19 IgG/IgM Test (RDT4, Nal Von Minden GmBH) and Finecare SARS-CoV2 Antibody test (RDT 9, Guangzhou Wondfo biotech).

Appraisal of test performance should also consider the influence of population prevalence, as it may change over time, geography and within different population groups. The potential risk of a test providing false positive results is crucial for release from lock-down of non-immune individuals. Among the 10 RDTs evaluated only three presented PPV and NPVs above 95 over a large window of population prevalence (RDT1, RDT 4, and RDT9).

These serological tests were able to independently diagnose COVID-19, especially in those with ≥2 weeks of symptoms, and could be used to triangulate unclear or false negative results from PCR and CT testing. They could also be used to monitor the status of medical and non-medical frontline workers and, over the longer term, to establish population level immunity as countries’ social restrictions ease. In the US (Santa Clara County, California) rapid antibody tests were used to evaluate the population prevalence of antibodies (ranging from 2.49-4.16) and helped authorities to understand that infection was far more widespread (55-fold) than indicated by the number of confirmed cases. These data are crucial to calibrate epidemic and mortality projections (29).

Among the three RDTs fulfilling the French health authorities’ criteria, only NG-Test IgG-IgM COVID-19 (NG-Biotech) might be considered a self-test since it includes all materials needed for self-puncture and capillary blood recovery. Nevertheless, we only authenticated this using serum, since its use has been previously established in capillary whole blood and our results in serum confirm those of the initial study (30). Namely, that this bedside fingerprick test confirmed infection in <15 minutes and could be performed by a medical practitioner without specialized training or a pathology laboratory (30).

Our study is limited in the following ways: (1) RT-PCR detection was based on upper respiratory tract specimens from patients with severe symptoms. None were asymptomatic patients (who did not access care). (2) Most study participants’ diagnoses were based on positive findings from an RT-PCR test using respiratory samples. Patients with negative RTPCR results but with chest imaging compatible with COVID-19 were not included. (3) Because the epidemic situation in France was very recent at the time of study, samples were collected during the acute phase of illness. Accordingly, we do not yet have sera from later stages to evaluate antibody persistence. (4) Only 10 out of more than 170 available RDTs have been evaluated.

The COVID-19 pandemic has revealed gaps in our diagnostic arsenal and is highlighting the essential role of serodiagnostics in public health response (31). With the use of carefully verified assays, appropriately designed serologic studies will help characterize transmission dynamics, refine disease burden estimates, diagnose suspected cases, and confirm clinically diagnosed patients without access to RT-PCR testing.

Though this assessment demonstrates varied analytical performance across a sample of current SARS-CoV-2 RDTs, they nevertheless hold real utility as tool for establishing population level exposure: many people have been exposed more than 3 weeks prior to antibody testing and would benefit from the nearly 100 sensitivity (in all tests evaluated) after 3 weeks’ time. However, highly sensitive (as early as 7 days) and specific tests are needed, both to achieve sufficiently high positive predictive values since population prevalence is often estimated to be low (≤5), and to be clinically useful as an initial diagnostic assay and a complement to direct RNA testing. Only three of the evaluated assays met the thresholds needed (sensitivity of >90 at 14 days after symptom appearance and >98 specificity).

Serological assays are simple, cheap, rapid, easy to interpret, and practical (can be stored at room temperature). They detect IgM, IgG, or both and can be performed directly at a patient’s bedside, at a general physician’s office, or when triaging in an emergency department, as most have been validated using whole blood.

## Data Availability

Data are available on request

